# The Association between Helicobacter pylori Infection and Triglyceride-Glucose (TyG) Index in US Adults: a restrospective cross-sectional study

**DOI:** 10.1101/2023.12.07.23299627

**Authors:** Wei Fu, Junlong zhao, GuoBin Chen, Linya Lyu, Yao Ding, Liang-Bi Xu

## Abstract

**Background:** The Triglyceride-glucose (TyG) index is an emerging marker for insulin resistance and metabolic syndrome. Helicobacter pylori, a bacterium associated with gastrointestinal diseases, may also influence metabolic risk profiles. This study aimed to investigate the relationship between the TyG index and Helicobacter pylori infection among a representative sample of adults.

**Methods:** A total of 9965 participants from the NHANES 1999-2000 cycle were involved from March 1, 1999, to December 1, 2000.A cross-sectional analysis was conducted on 3797 participants. The baseline characteristics according to the quartile of the TyG index were evaluated.Multivariate binary logistic regression models were constructed to analyze the independent effects of the TyG index on Helicobacter pylori infection. A two-piecewise binary logistic regression model was used to explore the non-linear relationship between the TyG index and Helicobacter pylori, with an inflection point identified. Subgroup analyses were performed to assess the trends of effect sizes across different variables including age, sex, glucose levels, body mass index (BMI), and chronic kidney disease (CKD).

**Results:** Multivariate analysis indicated a linear relationship between the TyG index and Helicobacter pylori infection, suggesting differential influence of the TyG index on Helicobacter pylori infection. Subgroup analysis demonstrated significant interactions only for a few variables, with all p-values for interaction below 0.05.

**Conclusions:** The study suggests a linear association between the TyG index and Helicobacter pylori infection.These findings have implications for understanding the metabolic influences on Helicobacter pylori infection and may guide targeted interventions for at-risk populations.

## INTRODUCTION

The interplay between metabolic disorders and infectious diseases has garnered considerable attention in recent years, paving the way for a better understanding of their shared pathophysiological mechanisms. The Triglyceride-glucose (TyG) index, a simple and cost-effective surrogate marker of insulin resistance, has been increasingly recognized for its role in predicting metabolic syndrome and cardiovascular diseases[1–3]. Insulin resistance, a hallmark of type 2 diabetes, has been implicated in the alteration of immune responses, potentially influencing the susceptibility to and severity of infectious diseases, including those caused by Helicobacter pylor[4–6].

Helicobacter pylori, a Gram-negative bacterium, colonizes the gastric mucosa of approximately half of the global population, leading to a range of gastrointestinal diseases such as gastritis, peptic ulcers, and gastric cancer[7–9].Beyond its established role in gastrointestinal pathology, H. pylori infection has been associated with extragastric manifestations, including cardiovascular and metabolic derangements, suggesting a broader impact on human health[10–12]. Despite the known associations between metabolic abnormalities and H. pylori infection, the direct role of the TyG index in relation to H. pylori infection remains poorly understood.

Research indicates that Helicobacter pylori infection is associated with the progression of atherosclerosis and is significantly and independently correlated with higher levels of LDL-C (low-density lipoprotein cholesterol) and lower levels of HDL-C (high-density lipoprotein cholesterol). However, Helicobacter pylori infection is not related to obesity-related parameters, glucose tolerance, or systolic blood pressure [13].There are also studies with opposing conclusions, which explored the relationship between Helicobacter pylori infection and fasting plasma glucose (FPG) levels in a non-diabetic population. The study found that Helicobacter pylori infection is an independent risk factor for increased FPG levels, and persistent infection can lead to increased levels of FPG and triglycerides/high-density lipoprotein (TG/HDL), which could be risk factors for diabetes[14].Recent research findings on the association between Helicobacter pylori infection and insulin resistance and metabolic syndrome have been contradictory. Therefore, a comprehensive meta-analysis was conducted, which included 22 studies involving 206,911 subjects. The results suggest that Helicobacter pylori infection may be related to metabolic syndrome and insulin resistance. Both case-control studies and cohort studies have shown this association[15].

Investigating this relationship is crucial, as it may elucidate potential metabolic pathways through which H. pylori influences disease and inform targeted interventions for at-risk populations. This study aimed to explore the relationship between the TyG index and H. pylori infection in a large cross-sectional adult population.To address this hypothesis, we conducted a comprehensive analysis, considering both linear and non-linear models, to delineate the nature of the association between the TyG index and H. pylori infection. Furthermore, we performed subgroup analyses to determine if age, sex, glucose levels, body mass index (BMI), and chronic kidney disease (CKD) modified the observed associations. The findings of this study are expected to contribute valuable insights into the metabolic factors influencing H. pylori infection, potentially offering a new perspective on the management and prevention strategies for this common infection.

## METHODS

### Study population

The National Center for Health Statistics (NCHS) is responsible for conducting the National Health and Nutrition Examination Survey (NHANES), which is an ongoing research project supported by the Centers for Disease Control and Prevention (CDC) [16]. This study has adopted a composite sampling technique, including multi-stage, stratified, and cluster probability sampling methods, to ensure that the sample adequately represents the U.S. population[17–19]. The study data are derived from the 1999-2000 NHANES. Participants in this cycle included those with data on Helicobacter pylori infection and Tyg index[20, 21].All procedures received approval from the CDC Ethics Review Board, and written informed consent was secured from all participants. Since the investigators had no access to identifying information, the data analyzed in this study were anonymized and publicly available on the NHANES website. Consequently, the 925th Hospital Review Board determined that this study qualified as “non-human subjects” research[20].

The inclusion criteria for this study were based on the NHANES 1999-2000 cycle, with a total of 9965 subjects participating. Exclusion criteria were applied, which included participants with missing data for H. pylori serology, participants with missing data for gastric diseases, and participants with missing data for covariates .

### Variables

The primary independent variable of interest in this study is the baseline measurement of the triglyceride-glucose (TyG) index. The triglyceride-glucose (TyG) index was calculated using the formula: TyG = ln[(fasting triglycerides (mg/dL) × fasting glucose (mg/dL)) / 2], where “ln” denotes the natural logarithm[21].The dependent variable is the presence of Helicobacter pylori infection, which is assessed using a dichotomous variable. In this study, standard ELISA thresholds were employed to classify participants as seropositive for Helicobacter pylori (optical density (OD) value ≥1.1) or seronegative (OD value <0.9). Equivocal OD values ranging from 0.9 to 1.1 were excluded to avoid the potential for confounding statistical results[22, 23].

The following variables were included in the fully-adjusted model based on the following criteria: demographic data, variables reported in previous literature to affect the TyG index or Helicobacter pylori, and variables based on clinical experiences. The fully-adjusted model included the following variables:Continuous variables obtained at baseline:age,serum C-reactive protein,Glucose,and Triglycerides.Categorical variables obtained at baseline: gender, education, race,high blood pressure, own housing, alcohol behavior,smoke behavior,BMI,Gastrointestinal illness, and CKD(Chronic kidney disease)[24–30].Patients with Chronic Kidney Disease (CKD) were defined as those with an estimated Glomerular Filtration Rate (eGFR) of less than 60 ml/min/1.73 m2 and/or proteinuria (dipstick results positive at or above 1+), demonstrating stable kidney function for at least three months prior to the study commencement[31].

### Statistical analysis

Categorical variables were expressed as frequencies and percentages. The differences among the TyG groups (quartiles) were analyzed using various statistical tests: the χ2 test for categorical variables, the t-test for normally distributed variables, and the Mann-Whitney U test for variables with skewed distributions[32].

The analysis was conducted in three stages. The first stage involved the application of multivariate binary logistic regression models, resulting in three distinct models: Model 1 without any covariate adjustments; Model 2 adjusted solely for sociodemographic data; and Model 3, which incorporated the covariates listed in Table 1. In the second stage, to address potential nonlinearity between the TyG index and Helicobacter pylori infection, a generalized additive model (GAM) and smooth curve fitting via penalized splines were utilized. Should nonlinearity be confirmed, a recursive algorithm would determine the inflection point, followed by the development of a two-piecewise binary logistic regression model on either side of this point. The final stage entailed subgroup analyses using stratified binary logistic regression models. Continuous variables were categorized based on clinical thresholds or tertiles prior to conducting an interaction test, and effect modification was assessed through the likelihood ratio test.

**Table 1:**
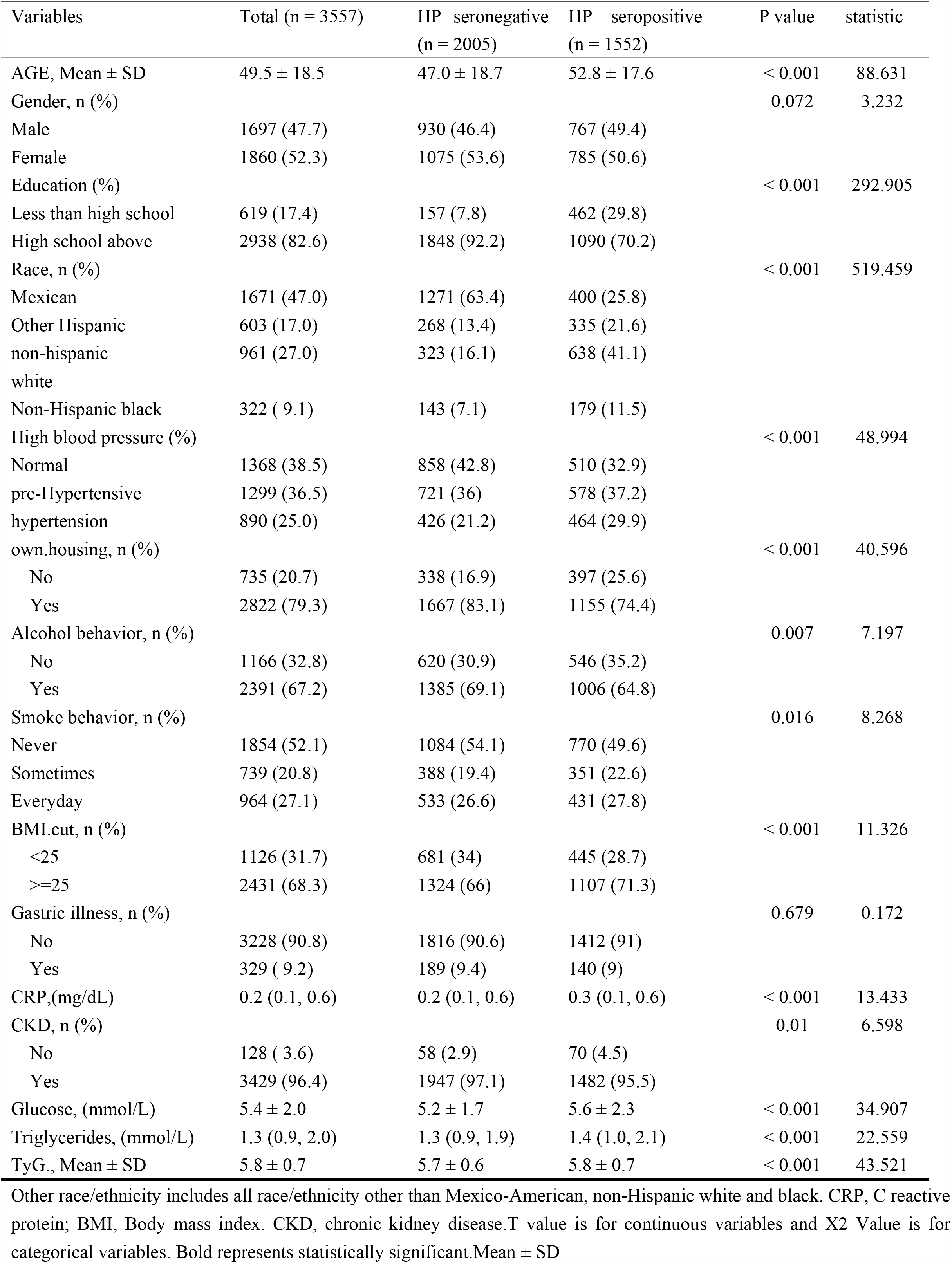
Baseline Characteristics of the Study Population: Comparison Between the Helicobacter pylori Seropositive and Seronegative Groups.

To confirm the robustness of the data analysis, a sensitivity analysis was performed. The TyG index was transformed into a categorical variable, and a P-value for trend was computed. The purpose of this analysis was to corroborate the findings obtained when the TyG index was treated as a continuous variable and to investigate potential nonlinearity.

Statistical analyses were conducted using the R software package version 4.2.2 (The R Foundation, http://www.R-project.org) and Free Statistics software version 1.9. A two-tailed test was employed, with a p-value less than 0.05 deemed indicative of statistical significance.

## RESULTS

### Baseline characteristics of selected participants

A total of 3797 participants were included in the final data analysis, as illustrated in Figure 1 (please refer to the flow chart). Table 1 presents the baseline characteristics of these selected participants according to the quartile of TyG index.

**figure 1.**
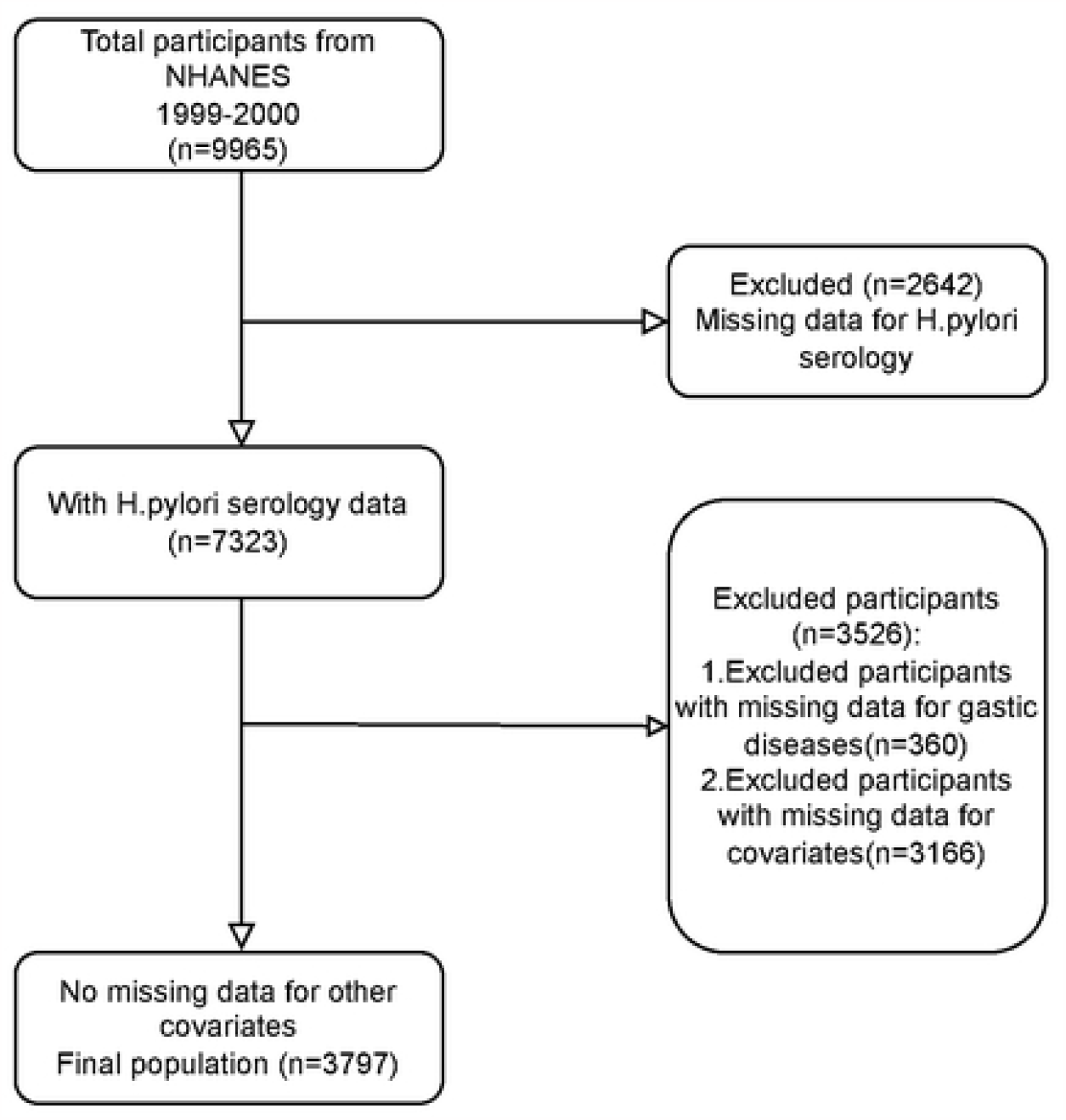

The average age of the study cohort was 41.8 years. The majority of participants were Mexicans (47%) with an education level higher than high school (>1) (82.6%), and there were individuals who never smoked (32.8%) or drank alcohol (52.1%). The mean TyG index was 5.8. Participants without HP infection had a significantly lower average TyG index, were younger, and had a lower BMI compared to those with HP infection. In terms of comorbidities, 25% of the study cohort had hypertension (Table 1). The group with HP infection had a higher incidence of hypertension and a history of cardiovascular diseases than the group without HP infection.

### Univariate analysis

Table 2 summarizes the univariate analysis of risk factors associated with Helicobacter pylori infection, reporting the disease risk in terms of OR and 95% CI. Age, Education, Race, own housing, High blood pressure, and TyG were significantly associated with Helicobacter pylori infection (p<0.001). Other factors, including Gastric illness and CRP, showed no significant association with Helicobacter pylori infection (Table 2).

**Table 2.** Univariate analysis of risk factor associated with Helicobacter pylori seropositivity.

| Variable | OR(95%CI) | P-value |
| --- | --- | --- |
| Age | 1.02 (1.01~1.02) | <0.001 |
| Sex |  |  |
| Male | Ref. |  |
| Female | 0.89 (0.78~1.01) | 0.072 |
| Education |  |  |
| Less than high school | Ref. |  |
| High school above | 0.2 (0.16~0.24) | <0.001 |
| Race, n (%) |  |  |
| Mexican | Ref. |  |
| Other Hispanic | 3.97 (3.26~4.83) | <0.001 |
| non-hispanic white | 6.28 (5.27~7.47) | <0.001 |
| Non-Hispanic black | 3.98 (3.11~5.09) | <0.001 |
| own.housing |  |  |
| No | Ref. |  |
| Yes | 1.71 (1.45~2.01) | <0.001 |
| Alcohol behavior |  |  |
| No | Ref. |  |
| Yes | 1.21 (1.05~1.4) | 0.007 |
| Smoke behavior, n (%) |  |  |
| Never | Ref. |  |
| Sometimes | 1.27 (1.07~1.51) | 0.006 |
| Everyday | 1.14 (0.97~1.33) | 0.106 |
| BMI, n (%) |  |  |
| <25 | Ref. |  |
| >=25 | 1.28 (1.11~1.48) | 0.001 |
| Gastric illness, n (%) |  |  |
| No | Ref. |  |
| Yes | 1.05 (0.83~1.32) | 0.679 |
| High blood pressure (%) |  |  |
| Normal | Ref. |  |
| pre-Hypertensive | 1.35 (1.16~1.57) | <0.001 |
| Hypertension | 1.83 (1.54~2.17) | <0.001 |
| CKD, n (%) |  |  |
| No | Ref. |  |
| Yes | 0.63 (0.44~0.9) | 0.011 |
| CRP | 1.06 (0.99~1.14) | 0.115 |
| TyG | 1.4 (1.26~1.55) | <0.001 |

To assess the independent effect of the TyG index on Helicobacter pylori, we developed three univariate and multivariate binary logistic regression models. Table 3 illustrates the odds ratio (OR) as the effect size and the corresponding 95% confidence intervals (CIs). In the unadjusted model (Model 0), the effect size represents the change in the risk of Helicobacter pylori infection associated with a one-unit difference in the TyG index. For instance, in the unadjusted model, a one-unit difference in the TyG index implies a 40% increase in the risk of Helicobacter pylori infection, with a 95% CI of [1.26, 1.55]. In the minimally adjusted model (Model 1), a one-unit increase in the TyG index is linked to a 27% rise in the risk of Helicobacter pylori infection, with a 95% CI of [1.14, 1.41]. In the fully adjusted model (Model 3), encompassing all covariates as displayed in Table 1, a one-unit increase in the TyG index is associated with an 18% increase in the risk of Helicobacter pylori infection, with a 95% CI of [1.04, 1.34].

**Table 3.** Multivariate logistical regression for TyG on Helicobacter pylori seropositivity..

| Variable | Model 0 |  | Model1 |  | Model2 |  | Model3 |  |
| --- | --- | --- | --- | --- | --- | --- | --- | --- |
|  | OR(95% CI) | P value | OR(95% CI) | P value | OR(95% CI) | P value | OR(95% CI) | P value |
| TyG | 1.4 (1.26~1.55) | <0.001 | 1.27 (1.14~1.41) | <0.001 | 1.17 (1.04~1.31) | 0.01 | 1.18 (1.04~1.34) | 0.008 |
| Q1(<5.203) | 1(Ref) |  | 1(Ref) |  | 1(Ref) |  | 1(Ref) |  |
| Q2(>=5.203,<5.553) | 1.15 (0.93~1.42) | 0.196 | 1 (0.81~1.25) | 0.986 | 0.97 (0.76~1.23) | 0.797 | 0.97 (0.76~1.24) | 0.824 |
| Q3(>=5.553,<5.86) | 1.21 (0.98~1.49) | 0.083 | 1.04 (0.84~1.29) | 0.723 | 0.93 (0.73~1.18) | 0.533 | 0.93 (0.73~1.19) | 0.577 |
| Q4(>=5.86,<6.249) | 1.41 (1.14~1.75) | 0.001 | 1.2 (0.96~1.49) | 0.108 | 1.15 (0.9~1.47) | 0.257 | 1.16 (0.9~1.49) | 0.243 |
| Q5(<=6.249) | 1.91 (1.55~2.36) | <0.001 | 1.54 (1.23~1.91) | <0.001 | 1.31 (1.03~1.68) | 0.031 | 1.33 (1.03~1.73) | 0.028 |
| P for trend |  | <0.001 |  | <0.001 |  | 0.01 |  | 0.009 |

Model 0: No covariates were adjusted.
Model 1: Adjusted for Age, Gender.
Model 2: Adjusted for Age, Gender, Edu, Race, Own.housing, Alcohol, Smoke.
Model 3: Adjusted for Age, Gender, Edu, Race, Own.housing, Alcohol, Smoke, BMI, Gastric.illness, HBP, CKD, CRP.

### Sensitivity analysis

For the sensitivity analysis, the TyG index was converted from a continuous variable to a categorical variable (five groups of the TyG index). The P-value for trend of the TyG index with categorical variables in the fully-adjusted model was 0.009, consistent with the results when the TyG index is treated as a continuous variable. Additionally, we observed that the trend of the effect size in different TyG index groups was not equidistant (Table 3).

This study analyzed whether there is a linear relationship between the TyG index and Helicobacter pylori infection . After adjusting for covariates, the smooth curve and the result of the generalized additive model indicated a linear relationship between the TyG index and Helicobacter pylori. Both binary logistic regression and two-piecewise binary logistic regression were used to fit the association, and the best-fit model was selected based on the P-value for the log likelihood ratio test. Since the P-value for the log likelihood ratio test was more than 0.05, the two-piecewise binary logistic regression was chosen to accurately represent the relationship between the TyG index and Helicobacter pylori. .

### Subgroup analysis

To observe the trend of effect sizes in different variables, we stratified the analysis by age, sex, glucose (glu), body mass index (bmi), and chronic kidney disease (CKD) (Figure 2). We found that, according to our a priori specifications, no interactions were observed, and the p-values for all interactions were greater than 0.05, indicating robust results.

**fig2.**
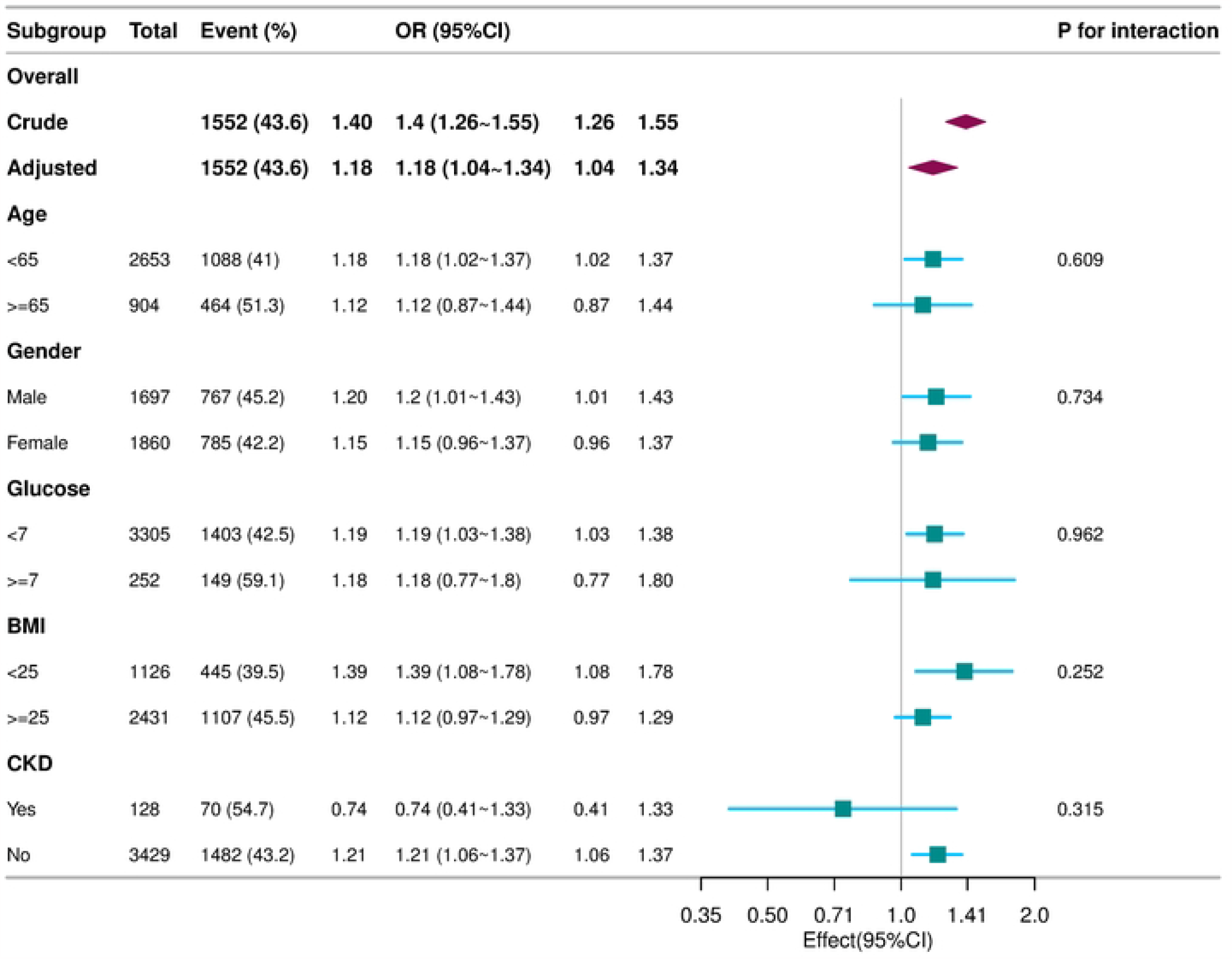

## DISUSSION

Our results clarify that there is a significant linear relationship between the triglyceride-glucose (TyG) index and Helicobacter pylori infection, which contributes to the growing body of literature on the metabolic effects of infectious diseases. Subgroup analysis shows that the relationship between the TyG index and Helicobacter pylori infection is modulated by factors such as age, gender, blood glucose levels, BMI, and chronic kidney disease (CKD), with no interactions found in any subgroup, leading to robust results.

These previous findings are consistent with reports that metabolic variables have differential impacts on infectious diseases, studying the role of metabolic health in the outcomes of H. pylori infection. Helicobacter pylori infection and eradication can selectively shape the metabolome and microbiome profiles of gastric lesions. However, how these different metabolic pathways, including isopentanoic acid, cholesterol, fatty acids, and phospholipids, specifically regulate the occurrence of gastric cancer or affect the host’s serum metabolism, thereby leading to the development of metabolic-related diseases, remains largely unknown[33].

Our results are consistent with previous studies suggesting a potential link between insulin resistance and the pathogenesis of H. pylori infection. You N, Chen Y et al have indicated that H. pylori infection is an independent risk factor for increased fasting plasma glucose (FPG) levels in non-diabetic (DM) individuals. Persistent H. pylori infection leads to elevated levels of FPG[34].

Helicobacter pylori infection (HPI) is diagnosed more frequently in patients with diabetes. Among them, insulin resistance in patients with type 1 diabetes (DMT1) is associated with the accumulation of advanced glycation end products (AGEs) in the skin and the progression of chronic complications[35].Song provided evidence that the eradication of Helicobacter pylori can improve glycemic control in patients with type 2 diabetes (T2DM), which indirectly reflects the interaction between Helicobacter pylori and diabetes[36].

It has also been discovered that diabetic patients with active Helicobacter pylori infection require higher intensity of glycemic treatment to achieve comparable blood sugar levels. Furthermore, the eradication of Helicobacter pylori has been shown to reduce A1C levels, thereby improving glycemic control[37].The results of a meta-analysis suggest that there is a possibility of metabolic syndrome and insulin resistance in individuals with Helicobacter pylori infection[15].

Research has found that the inflammatory response and inflammatory factors produced during HP (Helicobacter pylori) infection are important etiological factors for insulin resistance and metabolic syndrome (MS). The coexistence of long-term chronic inflammation and immune dysfunction may be precipitating factors for HP infection[38].

One possible mechanism is that inflammation associated with Helicobacter pylori may also lead to increased insulin resistance, thereby increasing the risk of diabetes in infected individuals. Additionally, the occurrence of skin diseases and ophthalmological conditions has been associated with this microorganism. In this context, this small review aims to collect major studies that link Helicobacter pylori infection with extragastric diseases and to explore the main mechanisms that may explain the role of Helicobacter pylori in these conditions[39].

Current research is focusing on the common pathogenic aspects between bacteria and insulin resistance or autoimmunity, the role of bacterial infections in cardiovascular risk, and whether infections worsen glycemic outcomes in diabetic patients. However, studies in this field still need to ultimately assess and interpret the potential relationship between Helicobacter pylori and diabetes. Some studies that agree have reached opposite conclusions[40].

The TyG index, as a marker of insulin resistance, may affect the immune system’s ability to respond to Helicobacter pylori, potentially influencing the efficiency of bacterial colonization or the host’s susceptibility to infection-related complications. The linear association we discovered adds to the complexity of understanding this relationship and emphasizes the need for further research to dissect the underlying biological mechanisms.

Our study highlights the importance of personalized approaches in managing H. pylori-infected patients by demonstrating that the association is not consistent across different levels of metabolic health.

The clinical significance of our study is substantial. Recognizing the linear dynamics between the TyG index and H. pylori infection may help clinicians identify individuals at high risk for infection or related complications. This could lead to earlier interventions, tailored treatment strategies, and potentially improved prognosis for patients with combined metabolic and infectious disease conditions. Our study includes a larger sample size compared to previous similar studies, enhancing the statistical power and generalizability of our findings. We specifically addressed the nonlinearity between variables in our study, allowing for a more comprehensive exploration of the association and providing a more accurate representation of the relationship. As an observational study, we employed strict statistical adjustment methods to minimize the impact of potential confounders, thus enhancing the internal validity of our results. We considered the TyG index as both a continuous and a categorical variable, reducing the chance of contingency in data analysis and improving the robustness of our findings.

However, our study has limitations. The generalizability of our findings is limited as the study subjects were restricted to the NHANES 1999-2000 cycle, which may limit the universality and extrapolation of our findings to broader populations. We excluded participants with missing data for H. pylori serology and covariates, so our results may not apply to these specific groups. The cross-sectional design precludes the establishment of causality, and the lack of longitudinal data limits our ability to determine the directionality of the observed associations. Moreover, the mechanisms by which H. pylori affects glycemic control are not yet certain and may require further exploration through biological experiments. Future longitudinal studies are needed to confirm our findings and explore the causal pathways involved.

In conclusion, our study indicates a linear relationship between the TyG index and H. pylori infection that is influenced by various metabolic and demographic factors. These insights pave the way for more detailed studies into the metabolic factors affecting H. pylori etiology and may ultimately lead to more effective patient management strategies.

## Data Availability

All data produced in the present study are available upon reasonable request to the authors
All data produced in the present work are contained in the manuscript
All data produced are available online

https://www.cdc.gov/nchs/nhanes/index.htm

## Author Contributions

FW contributed to the conceptualization and design of the study, data curation, and writing the original draft. ZJL played a significant role in methodology, formal analysis, and investigation. LLY DY CGB was involved in the validation, data curation, and analysis. provided resources, reviewed, and edited the manuscript. ZJL XLB, who is also the corresponding author, oversaw the project administration, supervised the study, and was responsible for funding acquisition. All authors have read and agreed to the published version of the manuscript.

## ACKNOWLEDGE

The authors thank all the staff members in our institution.We appreciate Dr. Jie Liu of the Department of Vascular and Endovascular Surgery, Chinese PLA General Hospital for statistics, study deign consultations and editing the manuscript.During the preparation of this work, we utilized AIGC for language polishing. Following the use of this service, the author reviewed and edited the content as necessary, taking full responsibility for the publication’s content.

